# Efficacy of Simultaneous Infusion of Tirofiban with Intravenous Thrombolysis in Patients with Acute Anterior Choroidal or Paramedian Pontine Infarction: TITACIPPI Study

**DOI:** 10.1101/2025.08.01.25332772

**Authors:** Moussa Toudou-Daouda, Djibril Soumah, Leila Bentamra, Nana-Rahamatou Aminou-Tassiou, Mohamed Baby, Yann Lhermitte, Tony Altarcha, Manvel Aghasaryan, Didier Smadja, Nicolas Chausson

## Abstract

**Background and Purpose:** Intravenous thrombolysis combined with tirofiban (IVT plus tirofiban) has been reported to be more effective than IVT alone in preventing early neurological deterioration (END) and improving 3-month functional outcomes in patients with ischemic stroke. This study aimed to assess the radiological efficacy and safety of simultaneous (or within 60 minutes) IVT plus tirofiban versus IVT alone specifically in patients with anterior choroidal infarction (ACI) or paramedian pontine infarction (PPI), two subtypes with a high risk of END.

**Methods:** We conducted a single-center, retrospective study including consecutive patients aged ≥18 years treated during two distinct periods: one with standard IVT alone and one with IVT plus tirofiban. The primary endpoint was ≥20% reduction in lesion volume on 24-hour diffusion-weighted imaging (DWI). Secondary endpoints included early clinical improvement at 24 hours and 7 days (or discharge), functional independence (modified Rankin Scale (mRS) scores ≤2 and ≤1) at 3 months, and END. Safety outcomes included symptomatic intracranial hemorrhage (sICH) and major systemic bleeding (MSB). A subgroup analysis compared clinical outcomes in patients with « lesion reversal » versus patients with no « lesion reversal ».

**Results:** A total of 58 patients were analyzed (30 IVT plus tirofiban; 28 IVT alone). Lesion reversal ≥20% was more frequently observed in the IVT plus tirofiban group (adjusted odds ratio, 8.58; 95% CI, 2.09–35.28; p=0.001). No significant differences were found between groups for END or 3-month functional outcomes (mRS ≤2 or ≤1). One non-fatal MSB occurred in the IVT alone group. No cases of sICH were observed in either group. Overall lesion reversal was significantly associated with better early and late outcomes, and with IVT plus tirofiban treatment.

**Discussion and Conclusions:** This study suggests that IVT plus tirofiban improves radiological outcomes and may be a promising therapeutic option in patients with ACI/PPI, without increasing the risk of bleeding compared to IVT alone.

**KEY MESSAGE:** **What is already known on this topic:** Branch atheromatous disease related ischemic strokes (BADRIS) carry a substantial risk of long-term disability because early neurological deterioration (END) is frequent even after intravenous thrombolysis (IVT). However, the role of IVT in preventing END in patients with BADRIS is not well established.

**What this study adds:** This single-center retrospective study provides real-world evidence that IVT plus tirofiban is associated with a lesion reversal ≥20% on 24-hour diffusion-weighted imaging (DWI) compared to IVT alone. The study also provides evidence that lesion reversal is associated with better early and late outcomes, and with IVT plus tirofiban treatment.

**How this study might affect research, practice or policy:** These findings support the use of IVT plus tirofiban in patients with BADRIS. This combination promotes lesion reversal, thereby improving functional outcomes and reducing the risk of END. Larger prospective multicenter randomized trials are needed to confirm these findings and to determine whether this strategy provides a functional benefit at 3 months.

## 1. Introduction

Paramedian pontine infarction (PPI) and anterior choroidal infarction (ACI) account for approximately 7% **[1]** and 2 to 8.3% **[2–4]** of all acute ischemic strokes (AIS), respectively, and are most often attributed to branch atheromatous disease (BAD) **[5]**. These AIS carry a substantial risk of long-term disability **[3, 4],** in part because early neurological deterioration (END) is frequent, between 25% and 60% of patients **[1–3, 6, 7],** even after intravenous thrombolysis (IVT) **[3, 7, 8]**. These results highlight the need to develop new therapeutic strategies for ACI/PPI.

Some preliminary studies conducted in Asia support a clinical benefit from administration of tirofiban within 24 hours following IVT in patients with ischemic strokes, without increased bleeding risk **[9–11]**. However, these studies involved heterogeneous stroke subtypes including both large-artery atherosclerosis, BAD and small-vessel disease. We believe that this strategy should preferably be evaluated as a priority in a homogeneous population of ischemic strokes most likely to be sensitive to this combined treatment, namely BAD. In addition, supporting evidence in favor of this strategy based on objective neuroimaging outcomes are lacking. Diffusion-weighted imaging (DWI) lesion volume at 24 hours has been proposed as a surrogate marker for long-term functional outcome in ischemic stroke and may serve as an early indicator of treatment efficacy **[12]**.

Thus, the present retrospective study aimed to compare the neuroradiological efficacy and safety of IVT plus simultaneous (or within 60 minutes) tirofiban versus IVT alone in patients with ACI/PPI, based on changes in lesion volume on DWI at 24 hours post-treatment.

## 2. Materials and Methods

### 2.1. Study Design

The TITACIPPI (Tirofiban with Intravenous Thrombolysis in acute Anterior Choroidal Infarction and Paramedian Pontine Infarction) study was a retrospective, single-centre study that compared a cohort of patients who were treated with IVT plus tirofiban (the intervention group) with a historical cohort who were treated with IVT alone (the control group). From the creation of our stroke unit in June 2012 until May 31, 2018, conventional IVT was the only therapeutic strategy used for acute ischemic stroke without arterial occlusion and in the absence of contraindications, specifically for ACI and PPI (control group). In June 2018, a change in our clinical practice was implemented whereby all patients admitted for acute ACI or PPI were treated with IVT plus tirofiban if there were no exclusion criteria (see below). In the intervention group, patients received tenecteplase (0.25mg/kg) or alteplase (0.9mg/kg), followed within 60 minutes by a bolus infusion of tirofiban (0.4μg/kg/min for 30 minutes), then a continuous intravenous infusion (0.1μg/kg/min for 48 hours). Tirofiban was then bridged with dual antiplatelet agents (aspirin plus clopidogrel or ticagrelor). This study was performed in accordance with the Strengthening the Reporting of Observational Studies in Epidemiology guidelines (STROBE) for reporting observational studies (Supplemental Material) and is registered with ClinicalTrials.gov, number NCT05733507.

### 2.2. Study Population

This study included patients with ACI/PPI, defined as follows **[8, 13]**: a) infarction with a diameter ≥20 mm in the perforating territory of anterior choroidal artery (lesion involved the posterior arm of the internal capsule and or posterior part of the corona radiata) or extending to the ventral pontine surface in the territory of paramedian pontine arteries; b) absence of large artery stenosis (>50%) or occlusion; c) no evidence of cardioembolism. Inclusion criteria were: (1) age ≥18 years; (2) National Institutes of Health Stroke Scale (NIHSS) score at admission ≥2; (3) for the intervention group, a time interval between IVT and intravenous tirofiban infusion ≤ 60 minutes, and (4) availability of a post-treatment brain magnetic resonance imaging (MRI) performed within 24-36 hours. Exclusion criteria were: (1) prestroke modified Rankin Scale (mRS) ≥3; (2) absence of MRI as initial imaging; (3) high-bleeding risk defined by severe cerebral microangiopathy (Fazekas score of 3/3 or old brain hemorrhage or ≥5 cerebral microbleeds on gradient echo MRI or recent major systemic hemorrhage; (4) aspirin administration within 24 hours after IVT; (5) absence of clear DWI-Fluid-attenuated inversion recovery (FLAIR) mismatch; and (6) patients with lenticulostriate artery infarctions, which usually affect a smaller portion of the pyramidal tract and are consequently associated with a lower risk of END.

### 2.3. Data Collection

Data were extracted from patients in the control group from March 1, 2014 to May 31, 2018 and from patients in the intervention group from June 1, 2018 to December 31, 2023. The following variables were collected for each participant: age at stroke onset, sex, vascular risk factors, history of stroke, history of myocardial infarction, NIHSS at admission, arterial territory (PPI or ACI), volume (ml) of the lesion on pre-treatment DWI, thrombolytic agent used (alteplase or tenecteplase), time between symptom onset and IVT (minutes), motor fluctuations before IVT (defined as transient clinical worsening), NIHSS at 24 hours post-IVT, END within 72 hours after IVT (defined as an increase of ≥4 points in total NIHSS or ≥2 points in motor function), lesion volume (mL) on post-treatment DWI at 24-36 hours, lesion “reversal” between pretreatment and post-treatment DWIs (see below), intracranial hemorrhage, systemic bleeding (urogenital, gastrointestinal, etc.), NIHSS at day 7 or discharge, and mRS at 3 months.

### 2.4. Efficacy and safety outcomes

The primary efficacy outcome of the study was the rate of “reversal” of the ischemic lesion at 24 hours. Lesion volumes were measured manually by multiplying the three dimensions of the hyperintense signal of the lesion on the pre-treatment and post-treatment DWI, based on a simplified ellipsoid volume formula (volume = [length × width × height]/2; ***Figure 1***), previously used in the estimation of intraparenchymal hematomas **[14]**. Two vascular neurologists (MTD and DSo), each with ≥10 years of clinical expertise in stroke imaging, consensually calculated all lesion volumes. To ensure unbiased analyses, the pretreatment and posttreatment DWI were analyzed several weeks after extraction of the baseline clinical data of patients and blinded to all clinical data. “Lesion reversal” was defined as a volume reduction of ≥20% on post-treatment DWI, based on the median percentage regression of the initial lesion observed after mechanical thrombectomy **[15]**. For each case of lesion reversal observed in DWI, the apparent diffusion coefficient (ADC) was used to confirm the reversal. However, the volumes were measured exclusively using DWI. Although manual measurement does not provide exact lesion volumes, the objective of this study was to determine whether the lesion volume had decreased by at least 20% between pre- and post-treatment DWIs, in order to classify patients as either « reversal » or « non-reversal ». As mentioned above, as the assessors were blinded to treatment allocation, differential misclassification is unlikely.

**Figure 1:**
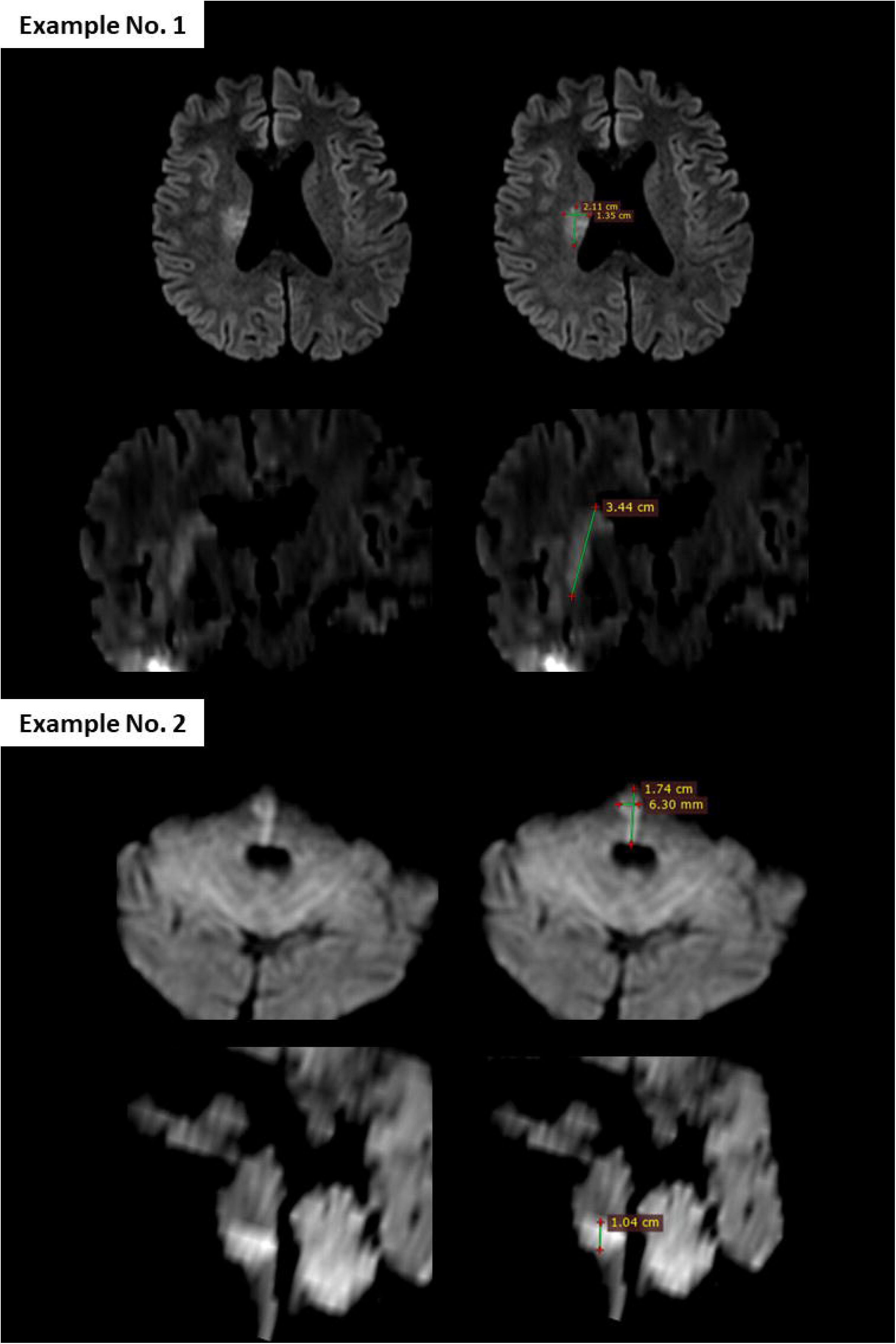
Diffusion-weighted imaging (DWI) from pretreatment brain MRI illustrating the method used to calculate lesion volume, based on the simplified ellipsoid formula: (Volume= (Length x Width x Height)/2). In example No. 1, the largest axial diameter was 2.11 cm, the perpendicular axial diameter was 1.35 cm, and the vertical diameter was 3.44 cm. The calculated lesion volume was (2.11 x 1.35 x 3.44)/2= 4.9 ml.

Secondary efficacy outcomes included delta-NIHSS at 24 hours (difference between initial and 24-hour NIHSS); a ≥2-point improvement in motor NIHSS items at 24 hours; and a major neurological improvement at 24 hours defined as ≥4-point overall NIHSS improvement. Additional outcomes included ≥30% and ≥40 lesion reversal on 24-hour DWI, ≥20 extension in lesion volume on 24-hour DWI, delta-NIHSS at 7 days or discharge, a ≥2-point improvement in motor NIHSS items at 7 days or discharge, END or clinical fluctuations within 72 hours post-IVT, mRS 0-2 and mRS 0-1 at 3 months.

Primary safety outcome was symptomatic intracranial hemorrhage (sICH), defined as intracranial hemorrhage causing neurological worsening with an increase of ≥4 points in total NIHSS or ≥2 points in one NIHSS category according to Heidelberg Bleeding Classification within 36 hours post-IVT **[16]** and major systemic bleeding (MSB), defined as a hemoglobin drop of ≥2 g/dL according to the International Society on Thrombosis and Haemostasis **[16]**. Secondary safety outcomes were minor bleeding (cerebral or systemic) and in-hospital mortality.

### 2.5. Ethics Statement

This study was conducted in accordance with the principles of the Declaration of Helsinki for Medical Research Involving Human Subjects, and the study protocol was approved by the ethics committee of Centre Hospitalier Sud-Francilien (Ref: 2022/0039). An information note, including the purpose of the study and a description of the data collected anonymously, was sent to each patient to inform them of their participation in the study, and the possibility to withdraw was offered to them. If no response was received within 30 days after the information notice was sent, the patient was deemed to have provided informed consent, in accordance with French regulations governing research involving human subjects.

### 2.6. Statistical Analysis

The Shapiro-Wilk test was used to assess normality for all quantitative variables. Baseline characteristics of the patients in the two groups were compared using Pearson’s Chi-squared test (or Fisher’s exact test where appropriate) for categorical variables and the Student’s t-test (or Mann-Whitney U test where appropriate) for continuous variables. The primary endpoint of the study was assessed using a binary logistic regression model with the treatment group as the explanatory variable. Secondary endpoints and safety measures of the study were also assessed using binary logistic regression models. Crude and adjusted odds ratios (ORs) were reported with corresponding 95% confidence intervals (CIs). Logistic regression models were also adjusted for key prognostic factors, including age, sex, baseline NIHSS score, lesion location (paramedian pontine artery vs. anterior choroidal artery), and baseline lesion volume measured on pre-treatment DWI. Patients with missing data for any of these variables were excluded from the adjusted analyses. The results of the lesion volume measurements on pre- and post-treatment DWI by both observers (MTD and DSo) were compared to determine interobserver agreement for lesion reversal threshold ≥20% (Cohen’s kappa coefficient). In addition, we compared clinical outcomes in patients with “reversion” with patients without “reversion”. Statistical analyses were performed using the SPSS statistical software package (IBM SPSS Statistics for Windows, Version 25.0. Armonk, NY: IBM Corp). P*-*values<0.05 were considered statistically significant.

## 3. Results

### 3.1. Baseline characteristics

During the study period from March 1, 2014, to December 31, 2023, a total of 136 patients were enrolled, of whom 59 met the study inclusion criteria, including 30 in the intervention group and 29 in the control group (***Figure 2***). After excluding one patient who withdrew from the study, 28 patients were finally included in the control group. The baseline characteristics of the patients were generally similar between the two groups except for initial lesion volume and initial NIHSS, which seem lower in the intervention group but without statistical significance (**Table 1**).

**Figure 2:**
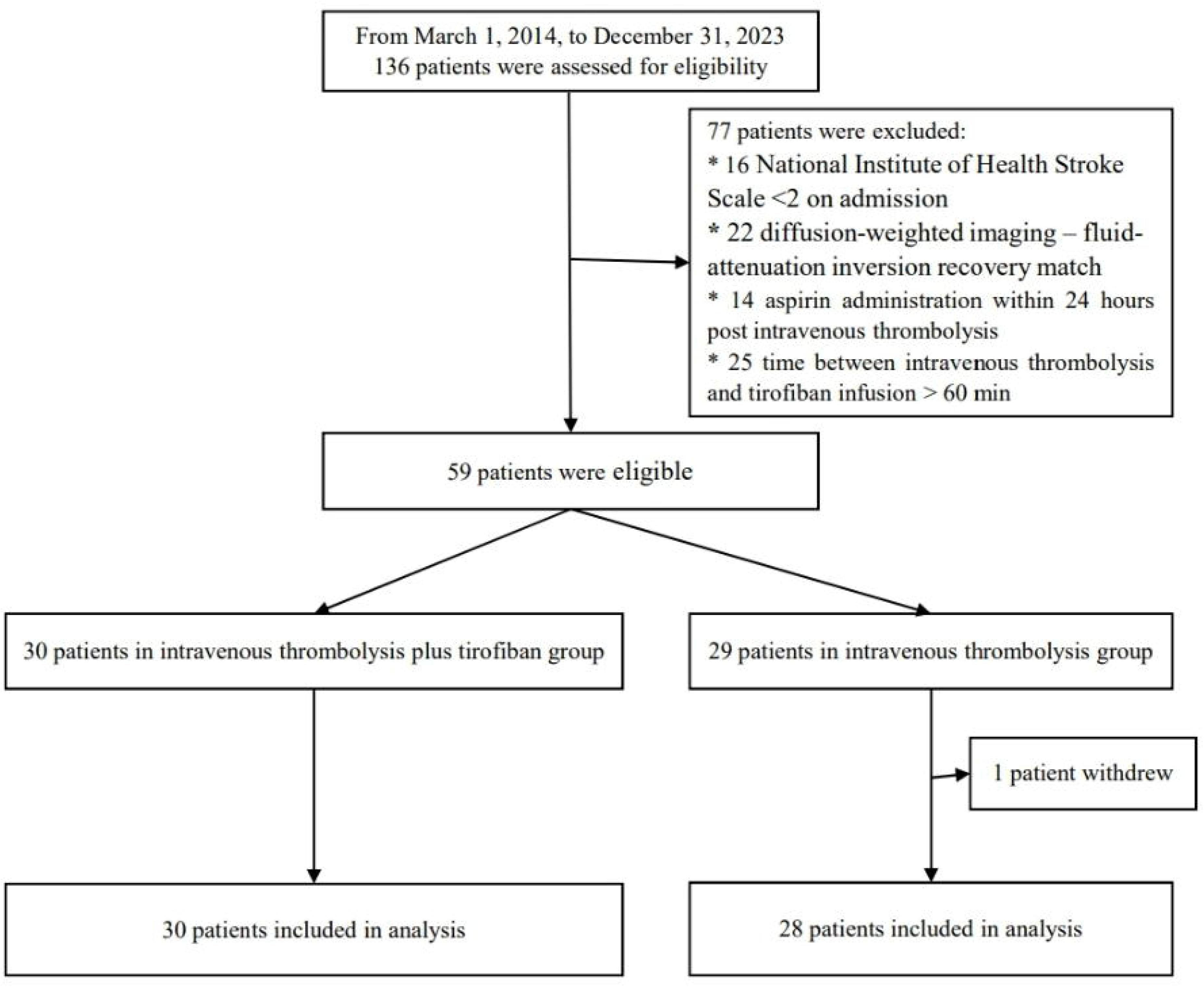
Flowchart of the study.

**Table 1:**
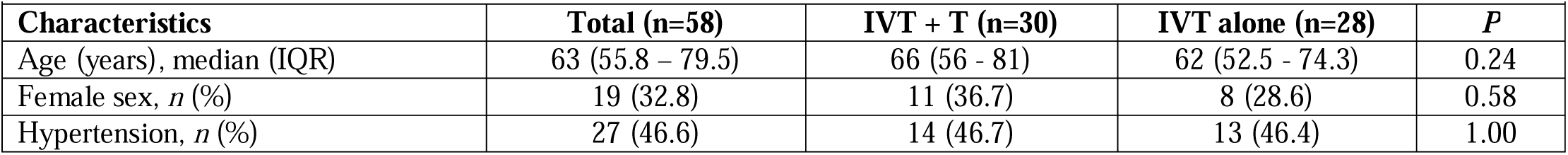
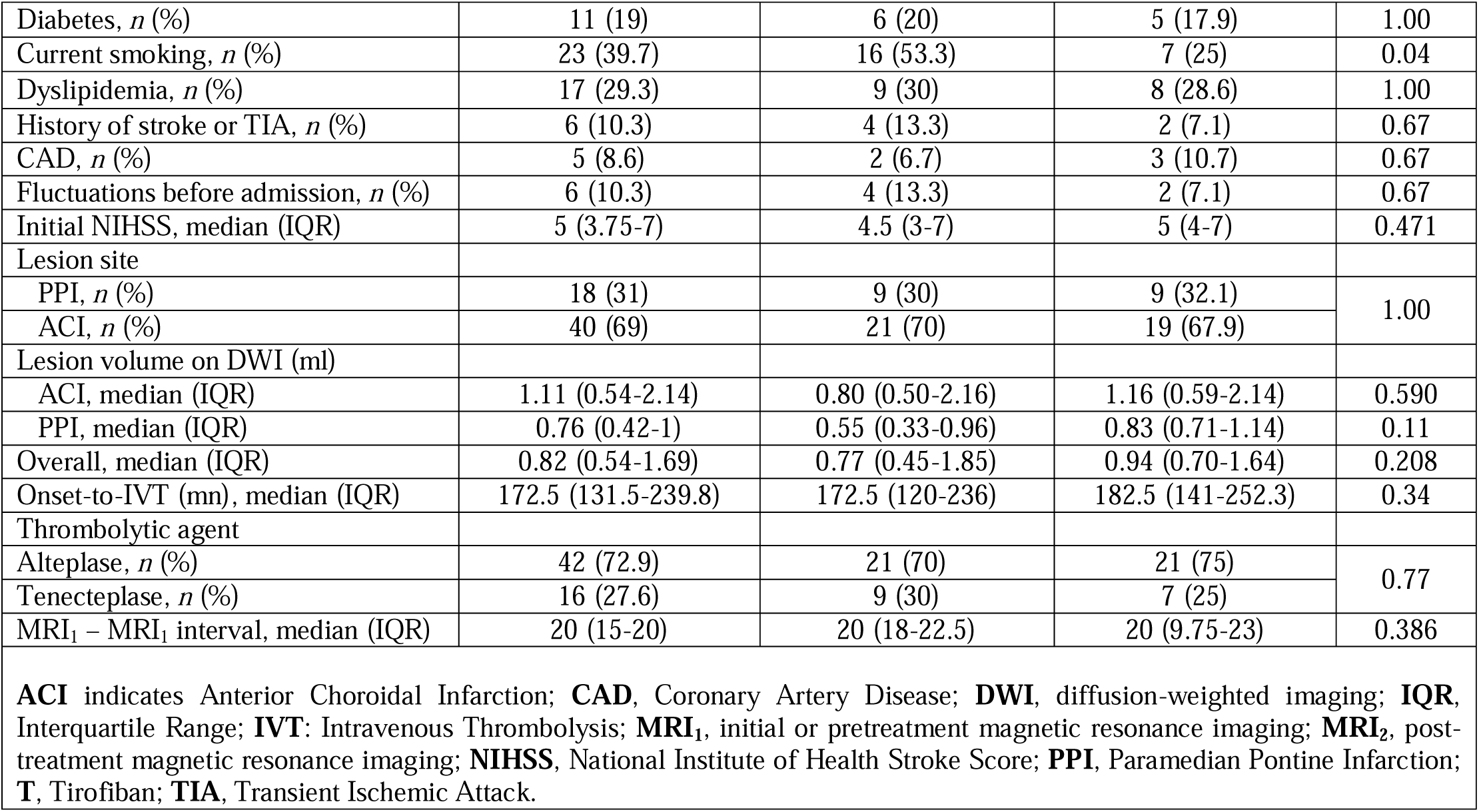
Baseline characteristics.

### 3.2. Primary Endpoint

Manual measurement found 19 cases of ≥20% DWI reversal at 24 hours out of 30 patients (63.3%) in the intervention group, and 6 out of 28 patients (21.4%) in the control group (adjusted OR, 8.58; 95% CI, 2.09 to 35.28; p=0.001) (**Table 2**). Intra-observer reproducibility was very good (K= 0.82; 95% CI, 0.50 to 1.00; p<0.001). ***Figure 3*** shows examples of lesion reversal ≥20%.

**Figure 3:**
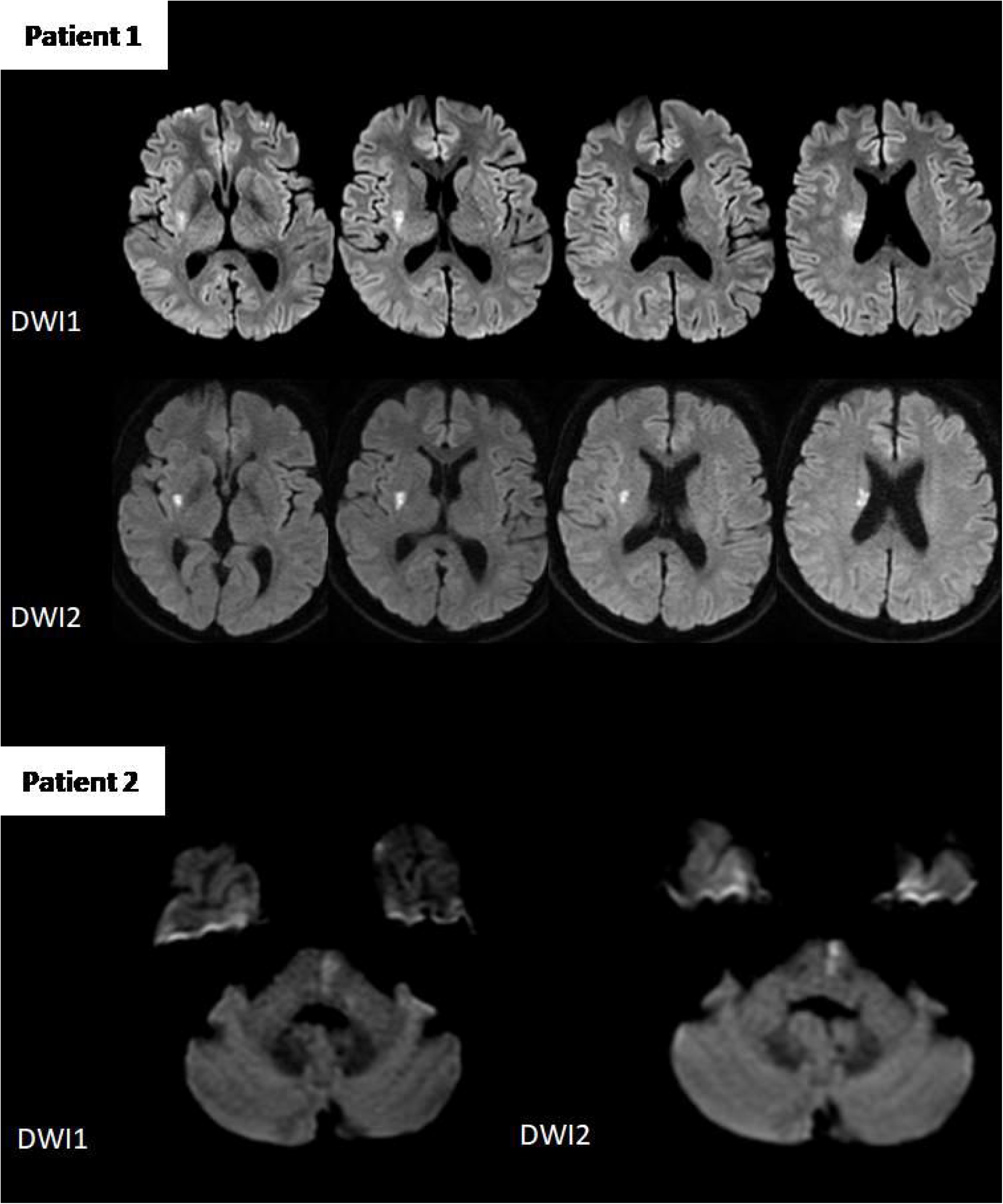
Lesion volume measurements from two patients in the IVT plus tirofiban group. Patient 1: Pretreatment DWI showed a lesion volume of 4.9 ml, which decreased to 1.6 ml on post-treatment DWI (67% reduction). Patient 2: Pretreatment lesion volume was 0.6 ml, reduced to 0.4 ml post-treatment (33% reduction).

**Table 2:** Study Outcomes.

| Table 2: Study Outcomes |  |  |  |  |  |  |
| --- | --- | --- | --- | --- | --- | --- |
| Outcomes | IVT + T<br>(n=30) | IVT alone<br>(n=28) | Crude OR<br>(95% CI) | *p* | Adjusted<br>OR<br>(95% CI) | *p* |
| **Primary efficacy endpoint** |  |  |  |  |  |  |
| Overall lesion reversal ≥20%, *n* | 19/30 | 6/28 (21.4) | 6.33 (1.97- | 0.002 | 8.58 (2.09- | 0.001 |
| (%) | (63.3) |  | 20.38) |  | 35.28) |  |
| ACI lesion reversal $\geq 20\%$, *n* (%) | 15/21<br>(71.4) | 6/19 (31.6) | 5.42 (1.40-20.97) | 0.014 | 5.25 (1.18-23.44) | 0.021 |
| PPI lesion reversal $\geq 20\%$, *n* (%) | 4/9 (44.4) | 0/9 (0) | 1.80 (1.01 -<br>3.23) | 0.041 | - | - |
| **Secondary efficacy endpoints** |  |  |  |  |  |  |
| Overall lesion extension $\geq 20\%$, *n* (%) | 9/30 (30) | 13/28 (46.4) | 2.02 (0.69-5.94) | 0.200 | 2.12 (0.57-7.85) | 0.260 |
| Overall lesion reversal $\geq 30\%$, *n* (%) | 17/30<br>(56.7) | 4/28 (14.3) | 7.85 (2.18-28.26) | 0.002 | 14.51 (2.76-76.23) | 0.002 |
| Overall lesion reversal $\geq 40\%$, *n* (%) | 15/30 (50) | 4/28 (14.3) | 6.00 (1.67-21.53) | 0.006 | 11.28 (2.11-60.32) | 0.005 |
| Delta-NIHSS at H24, median (IQR) | -2.5 ([-6]-<br>[0]) | -2.5 ([-4]-<br>[-1]) | 1.03 (0.90-1.18) | 0.667 | 1.10 (0.90-1.33) | 0.358 |
| Reduction in NIHSS $\geq 2$ at H24 | 20/30<br>(66.7) | 19/28 (67.9) | 1.06 (0.35-3.16) | 0.923 | 1.02 (0.32-3.30) | 0.971 |
| Reduction in NIHSS $\geq 4$ at H24 | 13/30<br>(43.3) | 7/28 (28.6) | 2.29 (0.75 -<br>7.03) | 0.146 | 4.63 (1.05-20.41) | 0.043 |
| Fluctuations after admission, *n* (%) | 3/30 (10) | 7/28 (25) | 3.00 (0.69-13.02) | 0.173 | 3.79 (0.78-18.45) | 0.099 |
| Delta-NIHSS at 7 days or discharge, median (IQR) | -3 ([-6.35]-<br>[0]) | -3 ([-5]-[-1]) | 1.05 (0.92-1.20) | 0.542 | 1.17 (0.95-1.42) | 0.136 |
| Reduction in NIHSS $\geq 2$ at 7 days or discharge | 22/30<br>(73.3) | 21/28 (75) | 1.09 (0.34-3.54) | 1.000 | 1.02 (0.30-3.41) | 0.998 |
| Reduction in NIHSS $\geq 4$ at 7 days or discharge | 14/30<br>(46.7) | 10/28 (35.7) | 1.58 (0.55-4.52) | 0.399 | 2.63 (0.64-10.77) | 0.178 |
| END, *n* (%) | 4/30 (13.3) | 8/28 (28.6) | 2.60 (0.69-9.87) | 0.160 | 3.31 (0.76-14.348) | 0.112 |
| mRS at 3 months |  |  |  |  |  |  |
| mRS 0-1, *n* (%) | 21/30 (70) | 15/28 (53.6) | 2.02 (0.69-5.94) | 0.200 | 2.91 (0.75-11.34) | 0.117 |
| mRS 0-2, *n* (%) | 23/30<br>(76.7) | 21/28 (75) | 1.10 (0.32-3.65) | 0.347 | 1.27 (0.34-4.74) | 0.347 |
| **Primary safety Endpoints** |  |  |  |  |  |  |
| sICH, *n* (%) | 0 | 0 | - | - | - | - |
| Major systemic bleedings, *n* (%) | 0 | 1/28 (3.6)<sup>a</sup> | 1.04 (0.97-1.17) | 0.483 | - | - |
| **Secondary safety endpoints** |  |  |  |  |  |  |
| Minor bleedings, *n* (%) | 9/30<br>(30)** | 1/28 (3.6)* | 11.57 (1.36-98.67) | 0.012 | 10.52 (1.10-100.73) | 0.041 |
| **ACI** indicates Anterior Choroidal Infarction; **END**, Early Neurological Deterioration; **IQR**, Interquartile Range; **IVT**: Intravenous Thrombolysis; **NIHSS**, National Institute of Health Stroke Score; **OR**, odds ratio; **PPI**, Paramedian Pontine Infarction; **sICH**, symptomatic Intracerebral Hemorrhage; **TIA**, Transient Ischemic Attack; **T**, Tirofiban.<br>* 1 minor intracerebral hemorrhage; ** 2 hematuria, 3 gingival bleedings, 2 epistaxis, 1 hematemesis, 1 rectal bleeding.<br><sup>a</sup> 1 metrorrhagia |  |  |  |  |  |  |

### 3.3. Secondary Endpoints

No significant clinical differences were observed at 24 hours between the two groups, with comparable overall delta-NIHSS scores (adjusted OR, 1.10; 95% CI, 0.90–1.33; p = 0.358). However, the proportion of patients achieving major neurological improvement at 24 hours was higher in the IVT plus tirofiban group compared to the IVT alone group (adjusted OR, 4.63; 95% CI, 1.05–20.41; p = 0.043) as well as achieving ≥30% (adjusted OR, 14.51; 95% CI, 2.76–76.23; p = 0.001) and ≥40% (adjusted OR, 11.28; 95% CI, 2.11–60.32; p = 0.005) lesion reversal on 24-hour DWI. No significant differences were found in clinical outcomes at 7 days or at discharge, nor in the proportion of patients achieving mRS ≤2 (adjusted OR, 1.27; 95% CI, 0.34–4.74; p = 0.347) or mRS ≤1 at 3 months (adjusted OR, 2.91; 95% CI, 0.75–11.34; p = 0.112).

### 3.4. Safety Endpoints

No cases of sICH occurred in both groups. MSB (metrorrhagia) occurred in 1 of 28 patients (3.6%) in the control group (non-fatal) and no patients in the intervention group (unadjusted OR, 1.04; 95% CI, 0.97 to 1.17; p = 0.483). Minor bleedings were more frequent in the intervention group compared to the control group, 9 vs. 1 respectively (**Table 2**). These included 2 cases of hematuria, 3 cases of gingival bleeding, 2 cases of epistaxis, 1 case of hematemesis and 1 case of rectal bleeding in the IVT plus tirofiban group. All of these events occurred within the first 24 hours of tirofiban infusion, and none resulted in discontinuation of the infusion early due to bleeding. However, no cases of asymptomatic intracranial hemorrhage were observed on follow-up MRI in the IVT plus tirofiban group, but one case was observed in the IVT alone group. No cases of in-hospital mortality occurred in both groups.

### 3.5. Subgroup analysis comparing patients with DWI reversal at 24 hours to those without reversal

A subgroup analysis was performed comparing patients who exhibited DWI lesion reversal at 24 hours to those who did not (**Table 3**). The two groups were clinically comparable, except for age: patients with lesion reversal were significantly older (median 67 years [IQR, 60–82]) compared to those without reversal (median 57 years [IQR, 49.5–73.5]; p = 0.021).

**Table 3:** Subgroup analysis comparing patients with DWI reversal at 24 hours to those without reversal.

|  |  |  |  |  |  |  |
| --- | --- | --- | --- | --- | --- | --- |
| **Outcomes** | **Reversal (n=25)** | **No reversal (n=33)** | **Crude OR (95% CI)** | ***p*** | **Adjusted OR (95% CI)** | ***p*** |
| Age (years), median (IQR) | 67 (60-82) | 57 (49.5-73.5) | 1.05 (1.01-1.08) | 0.021 | 1.07 (1.02-1.12) | 0.008 |
| Female sex, *n* (%) | 8/25 (32) | 11/33 (33.3) | 1.06 (0.35-3.22) | 1.000 |  |  |
| Hypertension, *n* (%) | 11/25 (44) | 16/33 (48.5) | 1.20 (0.42-3.40) | 0.795 |  |  |
| Diabetes, *n* (%) | 5/25 (20) | 6/33 (18.2) | 1.13 (0.30-4.21) | 1.000 |  |  |
| Current smoking, *n* (%) | 8/25 (32) | 15/33 (45.5) | 1.77 (0.60-5.24) | 0.417 |  |  |
| Dyslipidemia, *n* (%) | 7/25 (28) | 10/33 (30.3) | 1.12 (0.36-3.52) | 1.000 |  |  |
| History of stroke or TIA, *n* (%) | 3/25 (12) | 3/33 (9.1) | 1.36 (0.25-7.41) | 1.000 |  |  |
| CAD, *n* (%) | 2/25 (8) | 3/33 (9.1) | 1.15 (0.18-7.46) | 1.000 |  |  |
| Fluctuations before admission, *n* (%) | 2/25 (8) | 4/33 (12.1) | 1.59 (0.27-9.44) | 0.690 |  |  |
| Initial NIHSS, median (IQR) | 5 (3.5-7) | 5 (3.5-7) | 1.04 (0.89-1.22) | 0.594 |  |  |
| Onset-to-IVT (min), median (IQR) | 175 (128.5-263.5) | 170 (135-231.5) | 1.0 (0.99-1.01) | 0.756 |  |  |
| IVT plus tirofiban | 19/25 (76) | 11/33 (33.3) | 6.33 (1.97-20.38) | 0.002 | 5.99 (1.76-20.38) | 0.004 |
| IVT alone | 6/25 (24) | 22/33 (66.7) | 6.33 (1.97- | 0.002 |  |  |
|  |  |  | 20.38) |  |  |  |
| Alteplase use, *n* (%) | 17/25 (68) | 25/33 (75.8) | 1.47 (0.46-4.68) | 0.563 |  |  |
| Tenecteplase use, *n* (%) | 8/25 (32) | 8/33 (24.2) | 1.47 (0.46-4.68) | 0.563 |  |  |
| NIHSS at H24, median (IQR) | 0 (0-2) | 4 (1.5-5.5) | 1.63 (1.18-2.22) | 0.002 | 1.74 (1.25-2.41) | 0.001 |
| Reduction in NIHSS $\geq 2$ at H24 | 22/25 (88) | 17/33 (51.5) | 6.90 (1.73-27.60) | 0.005 | 16.18 (2.88-91.01) | 0.002 |
| Delta initial NIHSS - NIHSS at H24, median (IQR) | -4 ([-6]-[-2]) | -1 ([-3.5]-[1]) | 1.18 (1.01-1.37) | 0.037 | 1.73 (1.25-2.40) | 0.001 |
| NIHSS at 7 days or discharge, median (IQR) | 0 (0-0.5) | 3 (0.5-5.5) | 1.68 (1.17-2.41) | 0.005 | 1.79 (1.25-2.56) | 0.002 |
| Reduction in NIHSS $\geq 2$ at 7 days or discharge | 24/25 (96) | 19/33 (57.6) | 17.64 (2.13-146.76) | 0.001 | 53.85 (4.35-665.21) | 0.002 |
| Delta initial NIHSS - NIHSS at 7 days or discharge, median (IQR) | -4 ([-6.5]-[-3]) | -2 ([-5]-[1]) | 1.15 (1.0-1.33) | 0.056 | 1.79 (1.25-2.56) | 0.002 |
| END, *n* (%) | 2/25 (8) | 10/33 (30.3) | 5.0 (0.99-25.38) | 0.052 | 8.11 (1.36-48.46) | 0.022 |
| PPI, *n* (%) | 4/25 (16) | 14/33 (42.4) | 3.87 (1.08-13.81) | 0.045 |  |  |
| ACI, *n* (%) | 21/25 (84) | 19/33 (57.6) | 3.87 (1.08-13.81) | 0.045 |  |  |
| Lesion volume on DWI (ml) before treatment |  |  |  |  |  |  |
| ACI, median (IQR) | 1.16 (0.64-2.16) | 0.78 (0.54-2.14) | 1.0 (1.0-1.01) | 0.538 |  |  |
| PPI, median (IQR) | 0.47 (0.16-1.0) | 0.81 (0.52-1.0) | 1.0 (0.99-1.01) | 0.225 |  |  |
| Overall, median (IQR) | 1.06 (0.51-1.96) | 0.79 (0.55-1.34) | 1.0 (1.0-1.01) | 0.299 |  |  |
| Lesion volume on DWI (ml) after treatment |  |  |  |  |  |  |
| ACI, median (IQR) | 0.31 (0.13-0.46) | 1.50 (0.80-3.03) | 1.01 (1.01-1.02) | 0.003 |  |  |
| PPI, median (IQR) | 0.23 (0.01-0.74) | 1.07 (0.78-1.34) | 1.01 (1.0-1.01) | 0.061 |  |  |
| Overall, median (IQR) | 0.31 (0.01-0.46) | 1.15 (0.80-2.15) | 1.01 (1.01-1.02) | <0.001 |  |  |
| MRI<sub>1</sub> – MRI<sub>2</sub> interval | 20 (18.5-21.5) | 20 (10.5-23.5) | 1.01 (0.96-1.06) | 0.777 |  |  |
| mRS 0-1 at 3 months, *n* (%) | 20/25 (80) | 15/32 (46.9) | 4.53 (1.36-15.07) | 0.014 | 16.10 (2.45-105.64) | 0.004 |
| mRS 0-2 at 3 months, *n* (%) | 23/25 (92) | 20/32 (62.5) | 6.90 (1.38-34.60) | 0.013 | 23.75 (2.83-199.37) | 0.004 |

**ACI** indicates Anterior Choroidal Infarction; **END**, Early Neurological Deterioration; **IQR**, Interquartile Range; **IVT**: Intravenous Thrombolysis; **MRI<sub>1</sub>**, initial or pretreatment magnetic resonance imaging; **MRI<sub>2</sub>**, post-treatment magnetic resonance imaging; **NIHSS**, National Institute of Health Stroke Score; **OR**, odds ratio; **PPI**, Paramedian Pontine Infarction.

The IVT plus tirofiban strategy was used in 19 out of 25 patients (76%) who experienced lesion reversal, compared to only 11 out of 33 patients (33.3%) without lesion reversal (adjusted OR, 5.99; 95% CI, 1.76–20.38; **p = 0.004**). Conversely, only 6 out of 25 (24%) with lesion reversal where in the IVT group, compared to 22 out of 33 (66.7%) without lesion reversal.

Lesion reversal was strongly associated with clinical improvement at both 24 hours and at 7 days or discharge, across all evaluated parameters. At 24 hours, the median delta-NIHSS was -4 (IQR, -6 to -2) in the lesion reversal group versus -1 (IQR, -3.5 to 1) in the non-lesion reversal group (adjusted OR: 1.73; 95% CI, 1.25-2.56, **p = 0.002**). At 7 days or discharge, the median delta-NIHSS was -4 (IQR, -6.5 to -3) vs. -2 (IQR, -5 to 1), respectively (adjusted OR: 1.79; 95% CI, 1.25-2.56, **p = 0.002**). Lesion reversal also was associated with a reduction in END (adjusted OR: 8.11; 95% CI, 1.36-48.46, **p = 0.022**) and with achievement of mRS ≤1 (adjusted OR: 16.10; 95% CI, 2.45-105.64, **p = 0.004**) or mRS ≤2 (adjusted OR: 23.75; 95% CI, 2.83-199.37, **p = 0.004**).

## 4. Discussion

In this study of patients with acute ACI or PPI, simultaneous (or within 60 minutes) infusion of IVT plus tirofiban was associated with a higher likelihood of achieving lesion reversal ≥20% on post-treatment DWI compared to IVT alone. Regarding safety measures, no cases of sICH or in-hospital mortality were observed in either group. The combined treatment, IVT plus tirofiban, caused more minor bleedings, but none required discontinuation of tirofiban before the planned 48-hour duration, consistent with previous studies **[9, 10]**.

To our knowledge, this is the first study to evaluate the efficacy and safety of IVT plus tirofiban specifically in patients with ACI or PPI, and to propose an objective imaging surrogate marker for treatment response.

Imaging-based outcomes, such as early DWI lesion reversal, were shown to be more sensitive than clinical endpoints in detecting treatment effects in acute ischemic stroke, particularly in early-phase studies, and require smaller sample sizes to demonstrate efficacy **[15]**.

The potential benefit of combining IVT with tirofiban would be, first, to dissolve, with IVT, the thrombus formed on the ruptured microatheroma, and second to avoid its rapid reformation, through the antiplatelet activity of tirofiban. In our cohort, IVT plus tirofiban was significantly associated with major neurological improvement at 24 hours and appeared to reduce the risk of END, but not in a statistically significant way. However, IVT plus tirofiban was not associated with an improved mRS at 3 months. This may be explained by functional recovery achieved during rehabilitation in the IVT-alone group, which could offset early differences by 3 months. Alternatively, the lack of significant difference may be due to insufficient statistical power, given the very limited sample size, despite the substantial absolute difference in mRS 0–1 at 3 months rates (70% vs. 53.6%). On the other hand, our cohort showed a strong link between IVT plus tirofiban and lesion reversal, which reduces the risk of END and predicts major neurological improvement and three-month functional independence. Because the combined treatment IVT plus tirofiban seems to more often achieve DWI lesion reversal compared to IVT alone, we can therefore expect that this strategy can improve the final outcome and future large-scale prospective multicenter randomized trials are required to confirm this.

A major strength of our study is the inclusion of a strictly homogeneous etiological population, patients with BADRIS at high risk of END, mainly ACI or PPI. These infarct locations directly involve the pyramidal tract and are therefore particularly prone to disabling early motor deterioration, unlike lenticulostriate artery infarctions, which usually spare more pyramidal fibers and are less frequently associated with END. Because ACI and PPI carry a marked risk of worsening, they were specifically targeted in this therapeutic strategy. If effective in these high-risk subtypes, the approach may also benefit other clinically relevant BAD presentations. This approach avoids the heterogeneity present in studies that mix microatheroma, macroatheroma, and embolic strokes of undetermined source (ESUS) **[17]**. However, this also limits the generalizability of our results to other stroke subtypes.

Our study has several limitations. First, it is a retrospective, single-center, non-randomized study comparing patients treated in two different time periods. This may introduce potential major bias related to lesion reversal, possibly favoring the IVT plus tirofiban group. However, all imaging was performed on the same MRI scanner throughout the entire study period, using identical DWI acquisition parameters, thereby minimizing the risk of technical variability in lesion volume measurements. Second, the results should be interpreted with caution due to the small sample size, which may limit the statistical analyses power. In addition, systematic adjustment for confounders (e.g., propensity score matching) was not performed, and unmeasured confounding cannot be excluded. Third, the slightly smaller initial lesions in the intervention group may have favored better outcomes, despite no statistically significant difference.

In conclusion, the combination of IVT and tirofiban appears to increase the likelihood of early radiological improvement, which is associated with better neurological recovery in BAD-related ACI and PPI. On the other hand, the combination of IVT plus tirofiban was not associated with an improved mRS at 3 months due to limited sample size and possibly retrospective design comparing patients treated in two different time periods. However, the study results confirmed the safety profile of the combination of IVT plus tirofiban, because there was no increased risk of major bleeding compared to IVT alone. Finally, as our results are hypothesis-generating, future large-scale prospective multicenter randomized trials are required to confirm these findings and to determine whether this strategy provides a functional benefit at 3 months.

## Nonstandard Abbreviations and Acronyms

**ACI** indicates anterior choroidal infarction; **ADC**, apparent diffusion coefficient; **AIS**, acute ischemic strokes; **BAD**, branch atheromatous disease; **BADRIS**, branch atheromatous disease related ischemic stroke; **CI**, confidence intervals; **DWI**, diffusion weighted imaging; **END**, early neurological deterioration; **ESUS**, embolic strokes of undetermined source; **FLAIR**, Fluid-Attenuated Inversion Recovery; **IVT** intravenous thrombolysis; **MRI**, magnetic resonance imaging; **mRS**, modified Rankin Scale; **MSB**, major systemic bleeding; **NIHSS**, National Institutes of Health Stroke Scale; **OR**, odds ratios; **PPI**, paramedian pontine infarction; **sICH**, symptomatic intracranial hemorrhage.

## Acknowledgments

The sponsor was Centre Hospitalier Sud Francilien.

## Ethical approval

The study was conducted according to the principles of the Declaration of Helsinki and the study protocol was approved by the Ethics Committee of Centre Hospitalier Sud-Francilien (ref: 2022/0039).

## Informed consent

Each patient was informed of her/his participation in the study and was offered the possibility to withdraw.

## Trial Registration

This is registered with ClinicalTrials.gov, number NCT05733507.

## Guarantor

MTD.

## Data availability statement

The data that support the findings of this study are available on request from the corresponding author. The data are not publicly available due to privacy or ethical restrictions.

